# The age at onset of LRRK2 p.Gly2019Ser Parkinson’s disease across ancestries and countries of origin

**DOI:** 10.1101/2025.06.04.25327685

**Authors:** Theresa Lüth, Björn-Hergen Laabs, Sebastian Sendel, Inke R. König, Amke Caliebe, Alastair J. Noyce, Laurel A. Screven, Soraya Bardien, Matthew Farrer, Faycel Hentati, Christine Klein, Samia Ben Sassi, Joanne Trinh, Global Parkinson’s Genetics Program (GP2)

**Affiliations:** Institute of Neurogenetics, University of Lübeck, Lübeck, Germany; Institute of Medical Biometry and Statistics, University of Lübeck, Lübeck, Germany; Institute of Medical Informatics and Statistics, Kiel University and University Hospital Schleswig-Holstein, Kiel, Germany; Centre for Preventive Neurology, Wolfson Institute of Population Health, Queen Mary University of London, London, United Kingdom; National Institute on Aging, Bethesda, MD, USA; Division of Molecular Biology and Human Genetics, Faculty of Medicine and Health Sciences, Stellenbosch University and South African Medical Research Council/Stellenbosch University Genomics of Brain Disorders Research Unit, Cape Town, South Africa; Department of Neurology, University of Florida, Gainesville, FL, USA; Service de Neurologie, Institut National de Neurologie, La Rabta, Tunis, Tunisia; Faculty of Medicine of Tunis, 1006, Tunisia; Neurology’s Department, Mongi Ben Hmida National Institute of Neurology, Tunis, Tunisia

## Abstract

**Objectives:** The LRRK2 p.Gly2019Ser pathogenic variant has reduced penetrance and presents a wide range of age at onset (AAO) in patients with Parkinson’s disease (PD). We aim to elucidate differences in the cumulative incidence of LRRK2 p.Gly2019Ser*-*related PD (*LRRK2*-PD) between ancestries and countries.

**Methods:** We included N=922 unrelated LRRK2 p.Gly2019Ser variant carriers (affected: N=762, unaffected: N=160) from the Global Parkinson’s Genetics Program (GP2) in addition to cohorts recruited from the Israeli Ashkenazi Jewish and Tunisian Arab-Berber population. The p.Gly2019Ser variant was present in five ancestries: Ashkenazi Jewish (N=534), North African (N=223), European (N=132), Middle Eastern (N=19) and Latino and Indigenous people of the Americas (N=14). In addition to ancestry derived from the genetic data, we assessed the country of origin in our analysis. The Cox proportional-hazards model and Kaplan-Meier analysis were applied to examine differences in cumulative incidence. All analyses were adjusted for biological sex, and the outcome variable was AAO, including affected and unaffected variant carriers with right censoring for affection status, and all analysis were exploratory.

**Results:** The median AAO of *LRRK2*-PD was five years younger in the North African (HR=1.48, 95% CI: 1.18-1.86, p=7.0×10^−4^) compared to the European ancestry group. In contrast, the median AAO was five years older in the Ashkenazi Jewish (HR=0.61, 95% CI: 0.50-0.75, p=4.0×10^−6^) compared to the European ancestry group. Additionally, patients from Israel (HR=1.59, 95% CI: 1.30-1.39, p=4.0×10^−6^) and Tunisia (HR=2.57, 95% CI: 2.16-3.06, p<2.0×10^−16^) had a median 5-year and 10-year younger AAO compared to patients from the USA, respectively. Lastly, when focusing only on individuals with an Ashkenazi Jewish background, patients from Israel still had a younger AAO than those from the USA (HR=1.82, 95% CI: 1.48-2.24, p=1.5×10^−8^). Analogously, assessing only patients from the USA, the Ashkenazi Jewish ancestry group still had an older AAO than the European ancestry group (HR=0.51, 95% CI: 0.39-0.67, p=1.3×10^−6^).

**Discussion:** Our results provide evidence that a person’s genetic ancestry and country of origin are associated with the AAO of *LRRK2*-PD. This highlights the potential impact of both genetic and environmental factors on *LRRK2*-PD AAO.

## Introduction

Parkinson’s disease (PD) is the fastest-growing neurodegenerative disorder, currently affecting close to 12 million patients worldwide^1^. The etiology of PD is complex and multifactorial, and it is not yet completely understood. Environmental exposures, lifestyle, and genetics shape disease susceptibility. Approximately 15% of patients with PD have an underlying monogenic cause of the disease or carry a strong risk variant^2^. Among these, the LRRK2 (Leucine-rich repeat kinase 2) p.Gly2019Ser variant is the most common cause for autosomal dominant forms of PD. It is estimated that globally, 1-2% of all patients with PD carry the LRRK2 p.Gly2019Ser variant^2–4^. However, the frequency of the variant varies significantly, depending on the ancestral population. In Tunisian Arab Berbers, LRRK2 p.Gly2019Ser accounts for ∼30-40%^5–7^ of the PD cases, followed by Israeli Ashkenazi Jews at ∼18% of all PD cases^8–10^.

Although all p.Gly2019Ser variant carriers have the same genetic cause, they are not phenotypically homogeneous, which is evident in the varied age of onset (AAO) and disease severity across patients. In fact, not all persons who carry the p.Gly2019Ser variant will develop PD. Thus, reduced penetrance has been observed in many individuals carrying LRRK2 p.Gly2019Ser. Among unrelated mutation carriers, the risk of developing PD increases with age and has been initially estimated to be 28% at age 59 years, 51% at 69 years, and 74% at 79 years^11^ or even greater than 80% at 70 years^6^. In contrast, analyses in first or second-degree relatives of affected carriers using kin-cohort analyses presented lower estimates of the LRRK2 p.Gly2019Ser penetrance at ∼10-33% at 80 years^12, 13^. Thus, different studies assessed the penetrance of LRRK2 p.Gly2019Ser, with varying results depending on cohort compositions (e.g., families or unrelated mutation carriers) and statistical analysis approaches^14^. In addition, there is evidence for ancestral population-specific effects in LRRK2 p.Gly2019Ser-related PD (*LRRK2*-PD), as the penetrance is higher in the Arab-Berber population compared to USA Ashkenazi Jews and Europeans^5, 13^. Although a lower disease risk was also reported for p.Gly2019Ser variant carriers from North Africa ^15^. Concerning disease onset, no difference has been shown in AAO of *LRRK2*-PD patients from the USA or Tunisia; however, with a limited sample size^4^. In another study, Norwegian patients with *LRRK2*-PD had an older AAO compared to patients from the Tunisian Arab-Berber and Israeli Ashkenazi Jewish populations^16^.

A thorough assessment of the relationship between ancestry or the country of origin and *LRRK2*-PD onset has not been performed. Thus, we herein utilize data from the Global Parkinson’s Genetics Program (GP2)^17^ to assess the cumulative incidence in N=922 unrelated LRRK2 p.Gly2019Ser variant carriers from different ancestries and countries. The GP2 cohort includes patients from all over the world, with a particular focus on underrepresented populations. Utilizing resources like the GP2 dataset can help to overcome biases arising from limited statistical power and lack of non-European ancestry participants.

## Methods

### Study cohort

We included N=589 LRRK2 p.Gly2019Ser variant carriers from GP2, utilizing the genetic and clinical data of release 7 (https://gp2.org/)^17^. Additionally, we included LRRK2 p.Gly2019Ser variant carriers recruited from the Israeli Ashkenazi Jewish (N=127), as previously described^16^, and the Tunisian Arab Berber (N=217) populations.

The GP2 genotyping data, stored in PLINK format^18^, was utilized to extract the LRRK2 p.Gly2019Ser genotype (i.e., chr12:40340400 (hg38), rs34637584). The genotype data included in GP2 is obtained from the NeuroBooster array^19^ and was processed for quality control to encompass call rate pruning, evaluations for discordant sex, detect duplicates or related individuals (kinship, defaults: 0.0884/0.354 related/duplicated), and heterozygosity rates. Genotype imputation was performed through the TOPMed server^20^. All individuals were stratified into different ancestry groups by using the diverse reference panel based on prediction from the principal component analysis by GenoTools^21^, resulting in a classification of the individuals by GP2 into the following ancestry groups: African (AFR), African admixed (AAC), Ashkenazi Jewish (AJ), Latino and Indigenous people of the Americas (AMR), East Asian (EAS), European (EUR), South Asian (SAS), Central Asian (CAS), Middle Eastern (MDE), Finnish (FIN), and Complex Admixture (CAH) (**Supplementary Figure 1**). However, for the purposes of our study, we re-classified individuals from Tunisia (TUN) who were assigned to the MDE ancestry group to the North African ancestry group (NA). Subsequently, the genotype information was merged with the clinical metadata to obtain the affection status, age (i.e., age at sample collection), age of onset (if not available, age of diagnosis was used), biological sex and country of origin (i.e., three-letter country code). We only included ancestry groups with at least ten unrelated LRRK2 p.Gly2019Ser variant carriers with complete clinical data (i.e., AJ, AMR, EUR, MDE and NA). In total, we included N=922 variant carriers, consisting of N=762 patients affected by PD and N=160 unaffected variant carriers (**Table 1**).

**Table 1.** Demographics of LRRK2 p.Gly2019Ser variant carriers.

|  | *N* | *N* of men (%) | *N* of PD patients -<br>Mean AAO (*SD*) | *N* of unaffected carriers -<br>Mean AAE (*SD*) |
| --- | --- | --- | --- | --- |
| AJ | 534 | 273 (51.1%) | 395<br>59.81 years (11.32) | 139<br>61.90 years (11.35) |
| NA | 223 | 116 (52.0%) | 223<br>52.56 years (12.38) | - |
| EUR | 132 | 66 (50.0%) | 115<br>56.95 years (11.32) | 17<br>49.84 years (13.40) |
| MDE | 19 | 16 (84.2%) | 17<br>45.71 years (9.56) | 2<br>51.00 years (28.28) |
| AMR | 14 | 6 (42.9%) | 12<br>57.25 years (14.42) | 2<br>58.76 years (13.07) |
| **Total** | **922** | **477 (51.7%)** | **762**<br>**56.90 years (12.16)** | **160**<br>**60.44 years (12.29)** |

*N*=Number of individuals, AAE=Age at examination, AAO=Age at onset, PD=Parkinson's disease, AJ= Ashkenazi Jewish ancestry, AMR=Latino and Indigenous Americas populations, MDE=Middle Eastern ancestry, EUR=General European ancestry, NA=North African ancestry

In order to explore whether the observed difference in cumulative incidence across ancestries was specific to the group of patients with *LRRK2*-PD, we additionally included patients with PD who did not carry the LRRK2 p.Gly2019Ser variant (Non-*LRRK2*-PD). In total, we included N=10,892 patients with Non-*LRRK2*-PD that were from the AJ (N=1029) and EUR (N=9863) ancestry groups.

### Statistical analysis

All analyses and data visualization were conducted with the statistical software R (R v4.3.1)^22^. We utilized the Cox proportional-hazards model to assess the differences in cumulative incidence across different ancestries and countries and included affected and unaffected variant carriers. In the model, age (for unaffected variant carriers, censored) or AAO (for affected variant carriers), together with the affected/unaffected status, was the dependent variable, and ancestry and/or country of origin were the independent variables. Thus, event time was defined to be AAO with unaffected carriers being censored at their current age; and events were defined as affection with PD. The model was also adjusted for sex, given the unbalanced men-to-women ratios in some ancestry groups and the potential younger AAO of female *LRRK2*-PD patients^5^. The models were calculated with the R package *survival* (v3.5-7)^23^ and displayed as Forrest plots with the R package *forestmodel* (v 0.6.2, https://CRAN.R-project.org/package=forestmodel). Lastly, the age-associated cumulative incidences of the different ancestries or countries were visualized as Kaplan-Meier plots, and the survival curves were adjusted for sex. The Kaplan-Meier plots were visualized with the R package *survminer* (v0.4.9, https://CRAN.R-project.org/package=survminer). All analyses were exploratory, and p-values cannot be interpreted for significance.

## Results

In order to investigate the cumulative incidence of *LRRK2*-PD across ancestries and countries of origin, we assessed N=922 LRRK2 p.Gly2019Ser variant carriers (N=762 affected and N=160 unaffected). The variant carriers belonged to five different ancestries: EUR (N=132), AJ (N=534), AMR (N=14), MDE (N=19) and NA (N=223) (**Table 1**).

### LRRK2-PD cumulative incidence across different ancestries

We observed differences between the AAO of patients with *LRRK2*-PD across the ancestries (**Table 1, Supplementary Figure 2**). The AAO was the oldest in the AJ ancestry group (mean AAO (SD)=59.81 years (11.32)) and the youngest in the MDE group (mean AAO (SD)=45.71 years (9.56)). To statistically assess the relationship between ancestries and AAO of *LRRK2*-PD, we performed a Cox proportional-hazards analysis. We excluded the MDE (N=19) and AMR (N=14) ancestry groups for this analysis to counteract potential biases arising from too small sample sizes; still, an analogous analysis with all five ancestry groups is provided in **Supplementary Figure 2**.

In comparison to the EUR ancestry group, the median AAO was older in the AJ ancestry group (HR=0.61, 95% CI: 0.50-0.75, p=4.0×10^−6^) and younger in the NA ancestry group (HR=1.48, 95% CI: 1.18-1.86, p=7.0×10^−4^, **Figure 1A**). The sex-adjusted median age of survival without PD was ten years younger in the NA ancestry group (median age (95% CI)=54 years (52.9-56)) compared to the AJ ancestry group (median age (95% CI)=64 years (52.4-66)) and five years younger compared to the EUR ancestry group (median age (95% CI)=59 years (56-61)) (**Figure 1B**).

**Figure 1.**
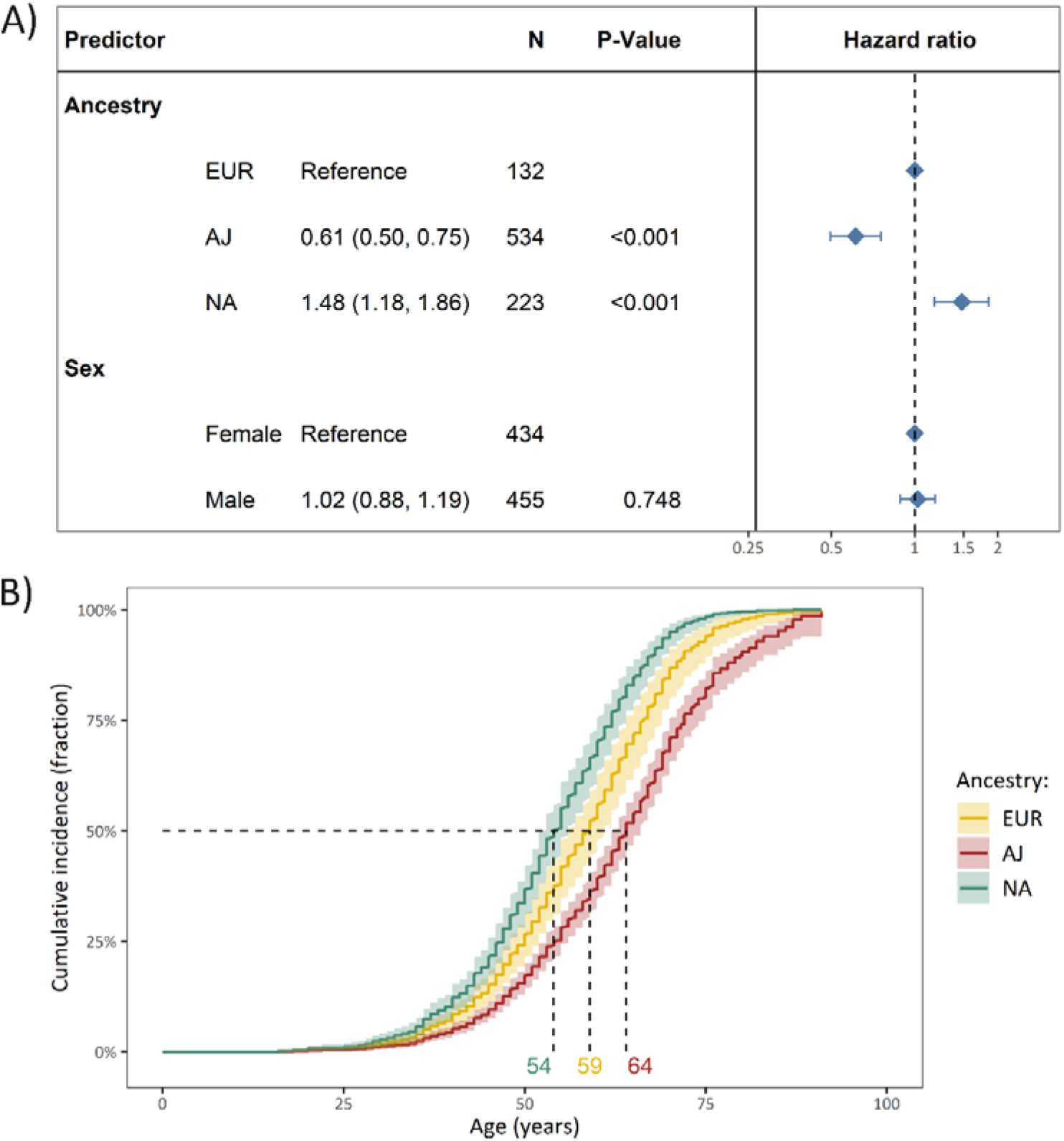
The difference in cumulative incidence of LRRK2 p.Gly2019Ser variant carriers from different genetic ancestries. **(A)** The forest plot indicates the difference in cumulative incidence of different ancestries where the hazard ratios, confidence intervals and *P*-values were derived from a Cox proportional-hazards model, adjusted for sex. The reference category of the assessed ancestries was set to European ancestry (EUR). Affected and unaffected LRRK2 p.Gly2019Ser variant carriers were included in the model and the outcome was age at onset or age at examination with right censoring of the affection status. (**B**) The Kaplan-Meier plot shows the adjusted curves with the respective confidence intervals, visualizing the Cox proportional-hazards model displayed above and the median age of disease onset. *N*=Number of individuals, Age=Age at examination or age at onset, PD=Parkinson’s disease, AJ=Ashkenazi Jewish ancestry, EUR=General European ancestry, NA=North African ancestry.

### LRRK2-PD cumulative incidence across different countries of origin

Next, we investigated the relationship between the country of origin and the cumulative incidence of *LRRK2*-PD. There were six countries with at least ten unrelated LRRK2 p.Gly2019Ser variant carriers: United States of America (USA, N=457), Tunisia (TUN, N=223), Israel (ISR, N=157), Spain (ESP, N=25), France (FRA, N=20), and India (IND, N=16). We observed differences between the median AAO of patients with *LRRK2*-PD across the countries of origin (**Supplementary Figure 3A**). To statistically assess these differences, we also performed a Cox proportional-hazards analysis. We included countries with at least 100 variant carriers for this analysis (i.e., USA, ISR and TUN). An analogous analysis with all six counties is provided in **Supplementary Figure 3**.

In comparison to the persons from the USA, the median AAO was younger among persons from ISR (HR=1.59, 95% CI: 1.30-1.39, p=4.0×10^−6^) and also younger among persons from TUN (HR=2.57, 95% CI: 2.16-3.06, p<2.0×10^−16^, **Figure 2A**). The sex-adjusted median age of survival without PD was ten years younger in variant carriers from TUN (median age (95% CI)=54 years (52-56)) compared to variant carriers from the USA (median age (95% CI)=64.1 years (63-66.3)) and six years younger compared to individuals from ISR (median age (95% CI)=60 years (57-62)) (**Figure 2B**).

**Figure 2.**
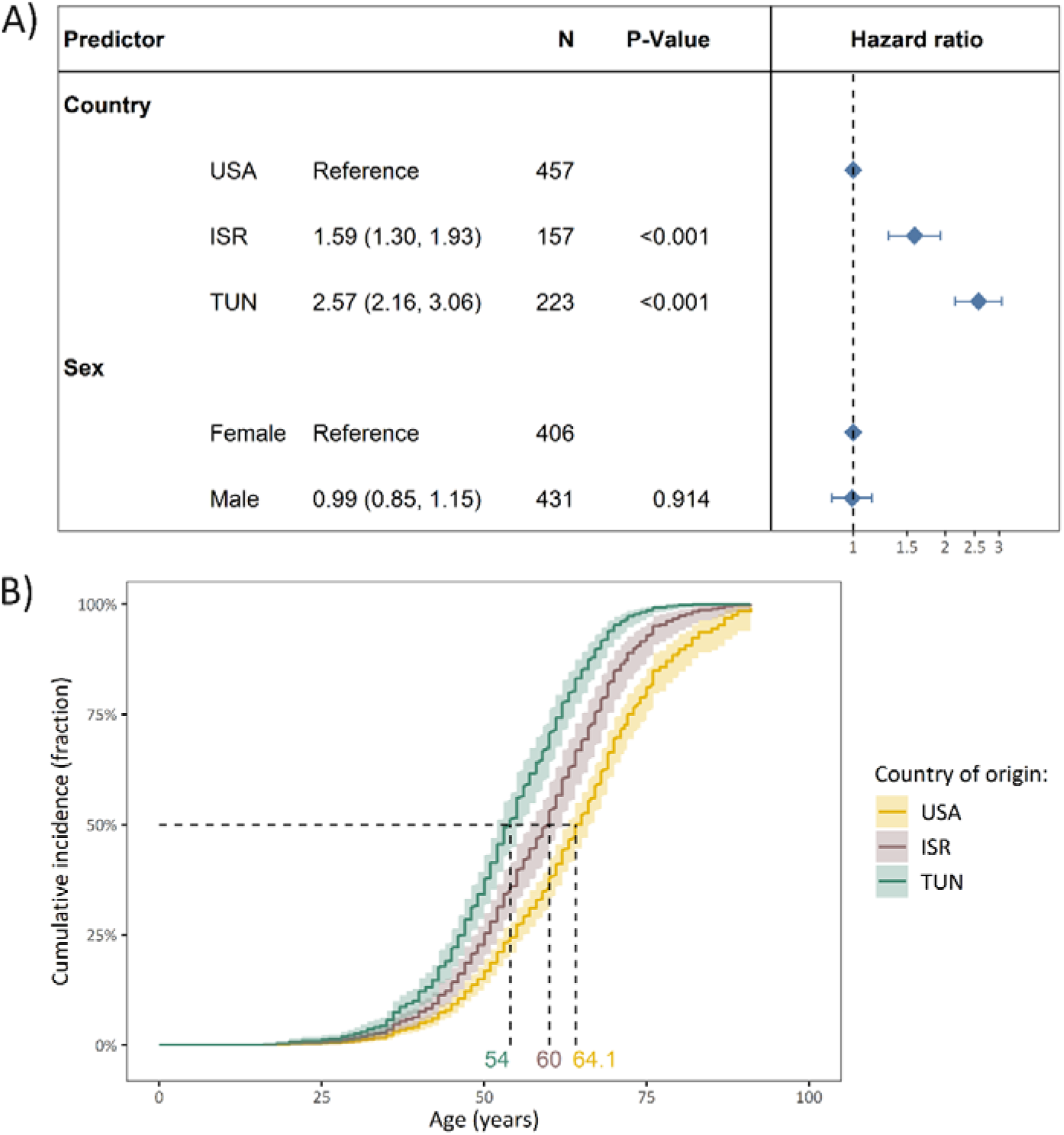
The difference in cumulative incidence of LRRK2 p.Gly2019Ser variant carriers from different countries of origin. **(A)** The forest plot indicates the difference in cumulative incidence from different countries where the hazard ratios and *P*-values were derived from a Cox proportional-hazards model, adjusted for sex. The reference category of the assessed countries was set to USA. Affected and unaffected LRRK2 p.Gly2019Ser variant carriers were included in the model and the outcome was age at onset or age at examination with right censoring of the affection status. (**B**) The Kaplan-Meier plot shows the adjusted curves with the respective confidence intervals, visualizing the Cox proportional-hazards model displayed above and the median age of disease onset. *N*=Number of individuals, Age=Age at examination or age at onset, PD=Parkinson’s disease, ISR=Israel, USA=United States of America, TUN=Tunisia.

### Ancestries and country of origin drive the difference in cumulative incidence

Next, we investigated if the difference in cumulative incidence of *LRRK2*-PD is driven by ancestry or country of origin. Thus, we focused on the ancestry group and country with the most included variant carriers, which were AJ and USA, respectively.

To test if the country of origin is associated with AAO beyond the genetic ancestry, we performed the Cox proportional-hazards analysis only in the AJ ancestry group. Within the AJ ancestry group, there were N=354 individuals from the USA and N=157 individuals from ISR. Notably, although all persons belonged to the same ancestry group, the median AAO was younger in patients from ISR (HR=1.82, 95% CI: 1.48-2.24, p=1.5×10^−8^). The median AAO was seven years younger in variant carriers from ISR (median age (95% CI)=60 years (58.7-62.1)) compared to variant carriers from the USA (median age (95% CI)=66.5 years (65-69)) (**Figure 3A**).

**Figure 3.**
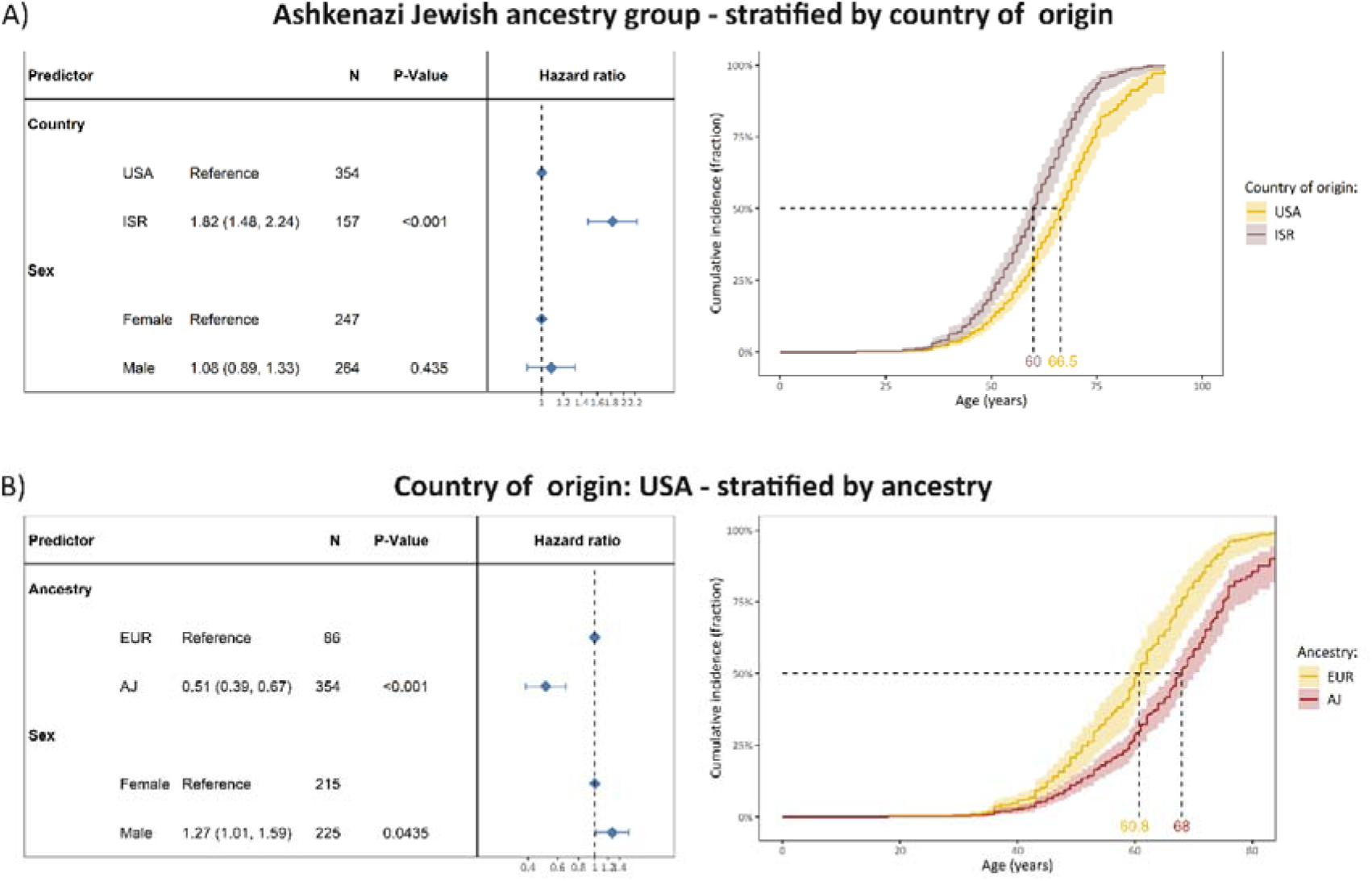
The difference in cumulative incidence of LRRK2 p.Gly2019Ser variant carriers from different countries of origin and ancestries. **(A)** The forest plot indicates the difference in cumulative incidence from different countries where the hazard ratios and *P*-values were derived from a Cox proportional-hazards model, adjusted for sex. The reference category of the assessed countries was set to USA. Affected and unaffected LRRK2 p.Gly2019Ser variant carriers were included in the model and the outcome was age at onset or age at examination with right censoring of the affection status. The Kaplan-Meier plot shows the adjusted curves and corresponding confidence intervals, visualizing the Cox proportional-hazards model displayed above and the median age at disease onset. All included mutation carriers were of Ashkenazi Jewish ancestry. **(B)** The forest plot indicates the difference in cumulative incidence from different ancestries where the hazard ratios and *P*-values were derived from a Cox proportional-hazards model, adjusted for sex. The reference category of the assessed countries was set to EUR. Affected and unaffected LRRK2 p.Gly2019Ser variant carriers were included in the model and the outcome was age at onset or age at examination with right censoring of the affection status. The Kaplan-Meier plot shows the adjusted curves and corresponding confidence intervals, visualizing the Cox proportional-hazards model displayed above and the median age at disease onset. All included mutation carriers originated from the USA. *N*=Number of individuals, Age=Age at examination or age at onset, PD=Parkinson’s disease, ISR=Israel, USA=United States of America, AJ=Ashkenazi Jewish ancestry, EUR=General European ancestry.

On the other hand, to test if ancestry is associated with AAO beyond the country of origin, we performed the Cox proportional-hazards analysis only in persons from the USA. Within the group of individuals from the USA, there were N=86 individuals with EUR ancestry and N=354 with AJ ancestry. Although all persons were from the same country, the median AAO was older in patients from the AJ ancestry group (HR=0.51, 95% CI: 0.39-0.67, p=1.3×10^−6^). The median AAO was approximately seven years older in variant carriers from the AJ ancestry group (median age (95% CI)=68 years (66-70.2)) compared to variant carriers from the EUR ancestry group (median age (95% CI)=60.8 years (59-64)) (**Figure 3B**).

Furthermore, we applied a Cox proportional hazards model with both country and ancestry as independent variables, including variant carriers from the EUR and AJ ancestry group as well as from ISR and the USA. Still, the countries of origin and ancestries remained associated with cumulative incidence (**Supplementary Figure 4**). In fact, individuals from the AJ ancestry group still had an older median AAO compared to individuals from the EUR one (HR=0.52, 95% CI: 0.40-0.68, p=2.3×10^−6^), and individuals from ISR still had a younger median AAO compared to individuals from the USA (HR=1.81, 95% CI: 1.47-2.23, p=1.8×10^−8^).

### Cumulative incidence across different ancestries of patients with PD that do not carry the LRRK2 p.Gly2019Ser variant

Lastly, we explored patients with PD who did not carry the LRRK2 p.Gly2019Ser variant (Non-*LRRK2*-PD). We focused our analysis on the ancestry groups relevant to this study. In total, we included N=10,892 patients with Non-*LRRK2*-PD from the AJ (N=1029) and EUR (N=9863) ancestry groups, where most of the patients were from the USA. Compared to patients with *LRRK2*-PD, the median AAO of patients with non-*LRRK2*-PD was approximately two years older in both the EUR (HR=0.82, 95% CI: 0.68-0.98, p=0.033) and AJ (HR=0.80, 95% CI: 0.72-0.90, p=2.7×10^−4^) ancestry group.

To assess whether the observed difference in cumulative incidence across ancestries was specific to the group of patients with *LRRK2*-PD, we analyzed patients without the p.Gly2019Ser variant. Analogous to the patients with *LRRK2*-PD, PD patients who did not carry the p.Gly2019Ser variant and belonged to the EUR ancestry group had a younger median AAO than those from the AJ ancestry group, albeit with a smaller effect size (HR=0.78, 95% CI: 0.73-0.83, p=1.4×10^−14^). There was a three-year difference in the median AAO of non-*LRRK2*-PD patients from the EUR (median AAO=60 years) and AJ (median AAO=63 years) ancestry groups (**Supplementary Figure 5**). The sample size of patients with non-*LRRK2*-PD from the NA ancestry group, Tunisia, or Israel was too small to allow for other ancestral or country-related comparisons.

## Discussion

In this study, we explored the relationship between genetic ancestry, country of origin and the cumulative incidence of *LRRK2*-PD. Utilizing one of the largest and most ancestral diverse study cohorts of LRRK2 p.Gly2019Ser variant carriers, we found evidence that both country of origin and genetic ancestry are associated with the cumulative incidence.

The median AAO was the oldest in the AJ ancestry group, followed by the EUR ancestry group and the youngest in the NA ancestry group. Thus, although all patients presumably had the same underlying monogenic cause of their disease, the ancestries’ different genetic backgrounds might impact the AAO. It is known that genetic modifiers are associated with disease risk and AAO in monogenic forms of PD. For *LRRK2*-PD in particular, although not yet replicated, *CORO1C* and *DNM3* have been nominated as genetic modifiers^24, 25^. Additionally, it has been demonstrated recently that a higher PD polygenic risk score is associated with a higher penetrance among LRRK2 p.Gly2019Ser variant carriers^26, 27^. Associations between ancestry and disease onset or disease risk have been observed in neurodegenerative diseases like Alzheimer’s Disease and amyotrophic lateral sclerosis^28, 29^.

We have previously assessed the implications of ancestry on the *LRRK2*-PD AAO and showed that Norwegians had a significantly later AAO compared to Tunisian Arab Berber and Israeli Ashkenazi Jewish populations^16^. Our data in this study could further highlight the large potential impact of genetic ancestry on the *LRRK2*-PD AAO. However, in our dataset, the EUR ancestry group had a ∼5 years younger median AAO compared to the AJ group. This discrepancy can result from the composition of individuals in the AJ ancestry group, as they were from the USA and Israel, whereas the previous study was limited to individuals from Israel. Additionally, individuals with EUR ancestry in this study here did not originate from Norway but are from various countries (e.g., USA, France or Spain).

Therefore, we next explored the relationship between countries and cumulative incidence. In contrast to ancestry, the country of origin is not derived from a person’s genetic background. The country of origin might and reflect similar environmental exposures or lifestyle preferences of persons living in the same geographical region. Still, it is important to note that the individual exposures can vary strongly between individuals within on country. In our dataset, individuals from the USA had the oldest median AAO. Individuals from TUN had a ∼10-year younger median AAO, and individuals from ISR had ∼5 years younger median AAO compared to individuals from the USA. Indeed, the importance of environmental and lifestyle factors for PD susceptibility and the association with *LRRK2*-PD AAO is known^30, 31^. Thus, it is reasonable that the country of origin is also associated with disease onset, although potential recruitment bias across the various countries might also impact this finding.

It is important to note that, in our study cohort, the ancestry group and country of origin are strongly correlated and subsequently not independent. To explore whether one or the other is driving the observed difference in cumulative incidences, we assessed the impact of the country within one ancestry group (i.e., AJ) and the impact of the ancestry within individuals from the same country (i.e., USA) (**Figure 3**). Importantly, the country and the ancestry were still associated with AAO. Thus, both might impact the AAO of *LRRK2*-PD. However, replicating this analysis within other countries and ancestries would be required to confirm this finding. Additionally, potential joint interaction effects between countries and genetic ancestry groups are possible, but given the strong correlation between country of origin and genetic ancestry group in our study, they cannot be investigated with this dataset.

Interestingly, analogous to the patients with *LRRK2*-PD, patients who did not carry the p.Gly2019Ser variant and belonged to the EUR ancestry group had a younger median AAO than those from the AJ ancestry group. However, the difference in non-*LRRK2*-PD was less pronounced, with only a three-year difference in median AAO. This observation might indicate that the ancestry-related difference in AAO was not limited to LRRK2 p.Gly2019Ser variant carriers, and thus, our findings might be of importance to different PD subtypes.

In this study, we did not observe that the biological sex was associated with *LRRK2*-PD AAO (**Figure 1-2**). Biological sex differences associated with *LRRK2*-PD AAO have been controversial in the literature. Females presented with a ∼5 years younger AAO in Tunisia^5^, and in Ashkenazi Jews, there was no such difference^13^. The difference in AAO of male and female patients with *LRRK2*-PD likely depends on the sampling ancestry group and country of origin, which should be explored further in future studies. For example, cultural and gender-related preferences of lifestyle factors might impact the difference in AAO of Tunisian male and female *LRRK2*-PD patients. One of the strongest protective lifestyle factors modifying PD risk and AAO is tobacco use, and the prevalence of smoking is substantially higher in Tunisian men compared to women^32^.

Besides genetics, lifestyle and environmental exposure, access to specialized health care and differences in socioeconomic factors are known to impact PD risk and AAO^33^, which likely contributed to the ancestry and country-specific AAO differences we observed. The main limitation of our study is that no information about environmental, lifestyle, socioeconomic factors, or access to specialized health care was available. Therefore, these factors could not be included in our statistical analysis, resulting in potential unaccounted biases. Thus, future studies should investigate the ancestry-specific effects of environment and lifestyle on *LRRK2*-PD AAO. Currently, lacking environmental and lifestyle data within GP2 restricts such analyses, but comprehensive efforts are underway to address these gaps.

There is also uncertainty about the variable country of origin, as there is the possibility that people have moved one or several times in their lifetime, resulting in potentially varying time spans that the individuals were exposed to respective environments. Additionally, there could be differences in patient assessments, recruitment and reported AAO between the different clinical sites within GP2 that we cannot account for. Furthermore, there was a large variability in the number of unaffected variant carriers among the ancestry groups. Survival analyses are also not adapted to case-control data and would benefit from prospective longitudinal studies. To exclude potential recruitment bias reflected in the unbalanced number of unaffected variant carriers and avoid biases from the interpretation of the statistical analysis, we repeated the Cox proportional-hazards model and only included affected patients. Still, the associations between ancestry, country and cumulative incidence remained unchanged (**Supplementary Figure 6**). Another limitation is that, in contrast to the AJ, EUR, and NA ancestry groups, the sample size of the AMR and MDE ancestries was too small to include them in the statistical analysis meaningfully. To achieve a more comprehensive understanding of *LRRK2*-PD, it would be valuable to enhance the representation of participants from these ancestries in research programs. Still, our study cohort presents the most extensive data set of LRRK2 p.Gly2019Ser variant carriers to explore the cumulative incidence across different ancestries and countries.

In conclusion, our results provide evidence that a person’s genetic ancestry and country of origin are associated with the AAO of LRRK2 p.Gly2019Ser-related PD. Future studies should aim to elucidate the underlying genetic, environmental and socioeconomic factors as well as potential gene-environment interactions leading to these observed AAO differences.

## Acknowledgment

This project was supported by the Global Parkinson’s Genetics Program (GP2; https://gp2.org). GP2 is funded by the Aligning Science Across Parkinson’s (ASAP) initiative and implemented by The Michael J. Fox Foundation for Parkinson’s Research (MJFF). For a complete list of GP2 members see doi.org/10.5281/zenodo.7904831.

## Data Availability

Data used in the preparation of this article were obtained from the Global Parkinson’s Genetics Program (GP2; https://gp2.org). Specifically, we used Tier 2 data from GP2 release 7 (https://doi.org/10.5281/zenodo.10962119). Tier 1 data can be accessed by completing a form on the Accelerating Medicines Partnership in Parkinson’s Disease (AMP®-PD) website (https://amp-pd.org/register-for-amp-pd). Tier 2 data access requires approval and a Data Use Agreement signed by your institution.

All code generated for this article, and the identifiers for all software programs and packages used, are available on GitHub [https://github.com/GP2code/AAO_LRRK2_pG2019S] and were given a persistent identifier via Zenodo [https://github.com/GP2code/AAO_LRRK2_pG2019S].

## Ethics Statement

We confirm that all necessary patient/participant consent has been obtained. The details of the IRB/oversight body that provided approval or exemption for the research described are given below:

1. Data used in the preparation of this article were obtained from the Global Parkinson’s Genetics Program (GP2; https://gp2.org). Specifically, we used Tier 2 data from GP2 release 7 (https://doi.org/10.5281/zenodo.10962119). Tier 1 data can be accessed by completing a form on the Accelerating Medicines Partnership in Parkinson’s Disease (AMP®-PD) website (https://amp-pd.org/register-for-amp-pd). Tier 2 data access requires approval and a Data Use Agreement signed by your institution.
2. Data from a cohort recruited from the Tunisian Arab Berber, and the study was approved by the ethics committee of the Mongi Ben Hmida National Institute of Neurology.
3. Data from a published cohort recruited from the Israel Ashkenazi Jewish population were included as described before by *Trinh J, Guella I, Farrer MJ. Disease Penetrance of Late-Onset Parkinsonism: A Meta-analysis. JAMA Neurol. 2014;71(12):1535–1539.* doi:10.1001/jamaneurol.2014.1909

## References

1. Collaborators GBDNSD. Global, regional, and national burden of disorders affecting the nervous system, 1990-2021: a systematic analysis for the Global Burden of Disease Study 2021. Lancet Neurol 2024;23:344–381.

2. Westenberger A, Skrahina V, Usnich T, et al. Relevance of genetic testing in the gene-targeted trial era: the Rostock Parkinson’s disease study. Brain 2024;147:2652–2667.

3. Deng H, Le W, Davidson AL, Xie W, Jankovic J. The LRRK2 I2012T, G2019S and I2020T mutations are not common in patients with essential tremor. Neurosci Lett 2006;407:97–100.

4. Ishihara L, Gibson RA, Warren L, et al. Screening for Lrrk2 G2019S and clinical comparison of Tunisian and North American Caucasian Parkinson’s disease families. Mov Disord 2007;22:55–61.

5. Trinh J, Amouri R, Duda JE, et al. Comparative study of Parkinson’s disease and leucine-rich repeat kinase 2 p.G2019S parkinsonism. Neurobiol Aging 2014;35:1125–1131.

6. Hulihan MM, Ishihara-Paul L, Kachergus J, et al. LRRK2 Gly2019Ser penetrance in Arab-Berber patients from Tunisia: a case-control genetic study. Lancet Neurol 2008;7:591–594.

7. Lesage S, Durr A, Tazir M, et al. LRRK2 G2019S as a cause of Parkinson’s disease in North African Arabs. N Engl J Med 2006;354:422–423.

8. Clark LN, Wang Y, Karlins E, et al. Frequency of LRRK2 mutations in early- and late-onset Parkinson disease. Neurology 2006;67:1786–1791.

9. Healy DG, Wood NW, Schapira AH. Test for LRRK2 mutations in patients with Parkinson’s disease. Pract Neurol 2008;8:381–385.

10. Ozelius LJ, Senthil G, Saunders-Pullman R, et al. LRRK2 G2019S as a cause of Parkinson’s disease in Ashkenazi Jews. N Engl J Med 2006;354:424–425.

11. Healy DG, Falchi M, O’Sullivan SS, et al. Phenotype, genotype, and worldwide genetic penetrance of LRRK2-associated Parkinson’s disease: a case-control study. Lancet Neurol 2008;7:583–590.

12. Goldwurm S, Tunesi S, Tesei S, et al. Kin-cohort analysis of LRRK2-G2019S penetrance in Parkinson’s disease. Mov Disord 2011;26:2144–2145.

13. Marder K, Wang Y, Alcalay RN, et al. Age-specific penetrance of LRRK2 G2019S in the Michael J. Fox Ashkenazi Jewish LRRK2 Consortium. Neurology 2015;85:89–95.

14. Trinh J, Schymanski EL, Smajic S, Kasten M, Sammler E, Grunewald A. Molecular mechanisms defining penetrance of LRRK2-associated Parkinson’s disease. Med Genet 2022;34:103–116.

15. Troiano AR, Elbaz A, Lohmann E, et al. Low disease risk in relatives of north african lrrk2 Parkinson disease patients. Neurology 2010;75:1118–1119.

16. Trinh J, Guella I, Farrer MJ. Disease penetrance of late-onset parkinsonism: a meta-analysis. JAMA Neurol 2014;71:1535–1539.

17. Global Parkinson’s Genetics P. GP2: The Global Parkinson’s Genetics Program. Mov Disord 2021;36:842–851.

18. Purcell S, Neale B, Todd-Brown K, et al. PLINK: a tool set for whole-genome association and population-based linkage analyses. Am J Hum Genet 2007;81:559–575.

19. Bandres-Ciga S, Faghri F, Majounie E, et al. NeuroBooster Array: A Genome-Wide Genotyping Platform to Study Neurological Disorders Across Diverse Populations. Mov Disord 2024;39:2039–2048.

20. Taliun D, Harris DN, Kessler MD, et al. Sequencing of 53,831 diverse genomes from the NHLBI TOPMed Program. Nature 2021;590:290–299.

21. Vitale D, Koretsky MJ, Kuznetsov N, et al. GenoTools: An Open-Source Python Package for Efficient Genotype Data Quality Control and Analysis. G3 (Bethesda) 2024.

22. R: A Language and Environment for Statistical Computing [computer program]. Version R version 4.3.1 (2023-06-16 ucrt) 2023.

23. Therneau TMG, P. M. Modeling Survival Data: Extending the Cox Model: Springer New York, NY, 2000.

24. Trinh J, Gustavsson EK, Vilarino-Guell C, et al. DNM3 and genetic modifiers of age of onset in LRRK2 Gly2019Ser parkinsonism: a genome-wide linkage and association study. Lancet Neurol 2016;15:1248–1256.

25. Lai D, Alipanahi B, Fontanillas P, et al. Genomewide Association Studies of LRRK2 Modifiers of Parkinson’s Disease. Ann Neurol 2021;90:76–88.

26. Kmiecik MJ, Micheletti S, Coker D, et al. Genetic analysis and natural history of Parkinson’s disease due to the LRRK2 G2019S variant. Brain 2024;147:1996–2008.

27. Iwaki H, Blauwendraat C, Makarious MB, et al. Penetrance of Parkinson’s Disease in LRRK2 p.G2019S Carriers Is Modified by a Polygenic Risk Score. Mov Disord 2020;35:774–780.

28. Chen HY, Panegyres PK. The Role of Ethnicity in Alzheimer’s Disease: Findings From The C-PATH Online Data Repository. J Alzheimers Dis 2016;51:515–523.

29. Roberts AL, Johnson NJ, Chen JT, Cudkowicz ME, Weisskopf MG. Race/ethnicity, socioeconomic status, and ALS mortality in the United States. Neurology 2016;87:2300–2308.

30. Luth T, Konig IR, Grunewald A, et al. Age at Onset of LRRK2 p.Gly2019Ser Is Related to Environmental and Lifestyle Factors. Mov Disord 2020;35:1854–1858.

31. Yahalom G, Rigbi A, Israeli-Korn S, et al. Age at Onset of Parkinson’s Disease Among Ashkenazi Jewish Patients: Contribution of Environmental Factors, LRRK2 p.G2019S and GBA p.N370S Mutations. J Parkinsons Dis 2020;10:1123–1132.

32. Nouira H, Ben Abdelaziz A, Rouis S, et al. Smoking behavior among students of health sciences at the university of Monastir (Tunisia). Tunis Med 2018;96:557–570.

33. Fernandes GC, Socal MP, Schuh AF, Rieder CR. Clinical and Epidemiological Factors Associated with Mortality in Parkinson’s Disease in a Brazilian Cohort. Parkinsons Dis 2015;2015:959304.

