## Supplementary for "The age at onset of LRRK2 p.Gly2019Ser Parkinson’s disease across ancestries and countries of origin"

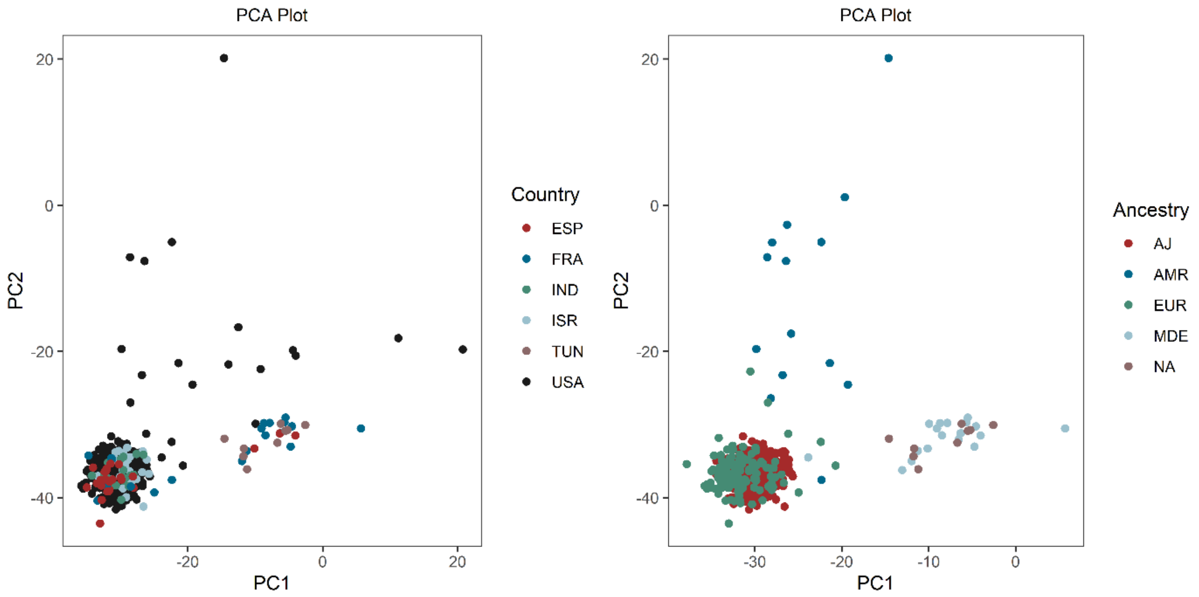


**Supplementary Figure 1. Principal component analysis (PCA).** The PCA plot displays the clustering of LRRK2 p.Gly2019Ser variant carriers of the GP2 dataset included in this study.

AJ=Ashkenazi Jewish ancestry, AMR=Latino and indigenous Americas populations, MDE=Middle Eastern ancestry, EUR=General European ancestry, NA=North African ancestry, ESP=Spain, FRA=France, IND=India, ISR=Israel, USA=United States of America, TUN=Tunisia.

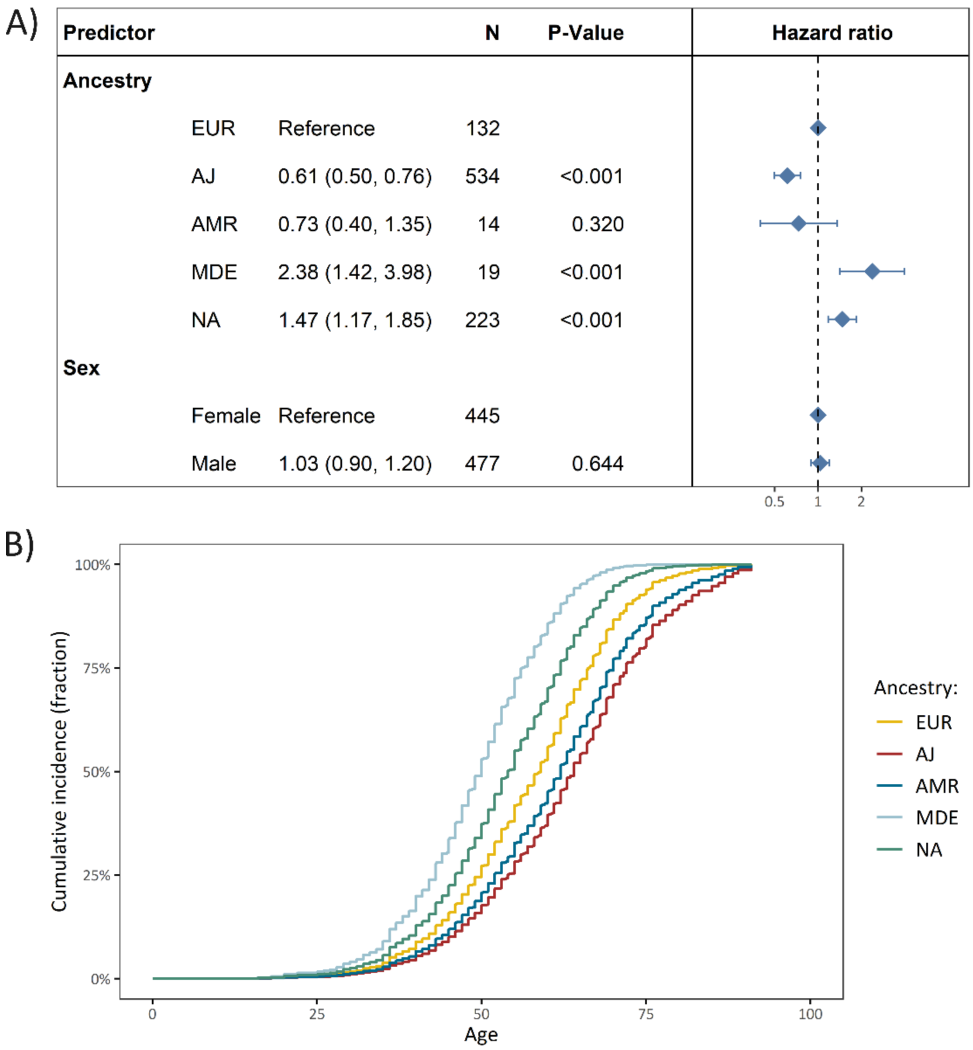


**Supplementary Figure 2. The difference in cumulative incidence of LRRK2 p.Gly2019Ser variant carriers from different genetic ancestries. (A)** The forest plot indicates the difference in cumulative incidence of different ancestries where the hazard ratios, confidence intervals and *P*-values were derived from a Cox proportional-hazards model, adjusted for sex. The reference category of the assessed ancestries was set to European ancestry (EUR). Affected and unaffected LRRK2 p.Gly2019Ser variant carriers were included in the model and the outcome was age at onset or age at examination with right censoring of the affection status. (**B**) The Kaplan-Meier plot shows the adjusted curves, visualizing the Cox proportional-hazards model displayed above.

*N*=Number of individuals, Age=Age at examination or age at onset, PD=Parkinson's disease, AJ=Ashkenazi Jewish ancestry, AMR=Latino and Indigenous Americas populations, MDE=Middle Eastern ancestry, EUR=General European ancestry, NA=North African ancestry.

A)

| Predictor | | | N | P-Value |
|---|---|---|---|---|
| **Country** | | | | |
| | USA | Reference | 457 | |
| | ESP | 2.42 (1.61, 3.64) | 25 | <0.001 |
| | FRA | 4.88 (3.05, 7.80) | 20 | <0.001 |
| | IND | 0.47 (0.21, 1.07) | 16 | 0.0709 |
| | ISR | 1.59 (1.30, 1.93) | 157 | <0.001 |
| | TUN | 2.56 (2.15, 3.05) | 223 | <0.001 |
| **Sex** | | | | |
| | Female | Reference | 429 | |
| | Male | 1.00 (0.86, 1.16) | 469 | 0.9908 |

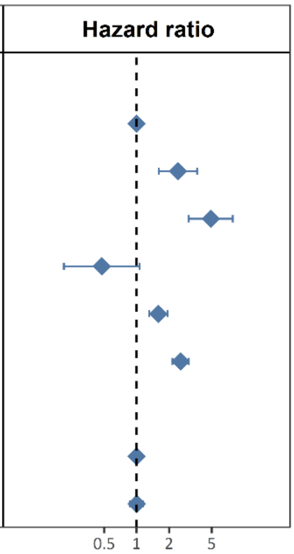


B)

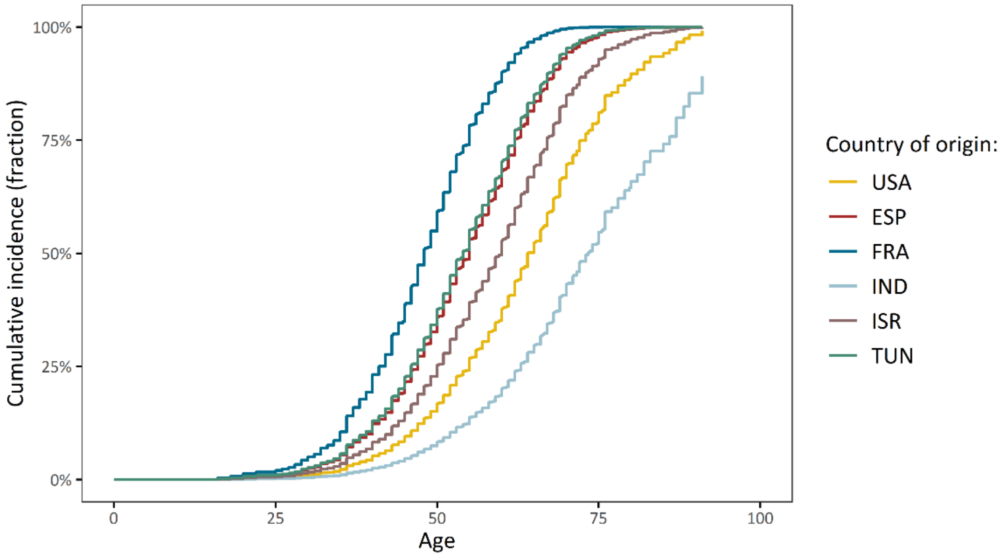


**Supplementary figure 3. The difference in cumulative incidence of LRRK2 p.Gly2019Ser variant carriers from different countries of origin. (A)** The forest plot indicates the difference in cumulative incidence from different countries where the hazard ratios and *P*-values were derived from a Cox proportional-hazards model, adjusted for sex. The reference category of the assessed countries was set to USA. Affected and unaffected LRRK2 p.Gly2019Ser variant carriers were included in the model and the outcome was age at onset or age at examination with right censoring of the affection status. (**B**) The Kaplan-Meier plot shows the adjusted curves, visualizing the Cox proportional-hazards model displayed above.
*N*=Number of individuals, Age=Age at examination or age at onset, PD=Parkinson's disease, ESP=Spain, FRA=France, IND=India, ISR=Israel, USA=United States of America, TUN=Tunisia.

| Predictor | | | N | P-Value |
|---|---|---|---|---|
| **Ancestry** | | | | |
| | EUR | Reference | 86 | |
| | AJ | 0.52 (0.40, 0.68) | 511 | <0.001 |
| **Country** | | | | |
| | USA | Reference | 440 | |
| | ISR | 1.81 (1.47, 2.23) | 157 | <0.001 |
| **Sex** | | | | |
| | Female | Reference | 290 | |
| | Male | 1.11 (0.92, 1.34) | 307 | 0.266 |

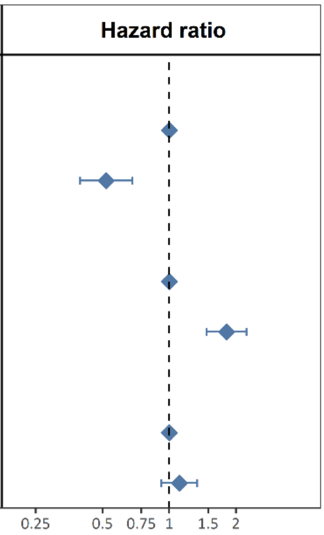


**Supplementary Figure 4. The difference in cumulative incidence of LRRK2 p.Gly2019Ser variant carriers from different genetic ancestries and countries.** The forest plot indicates the difference in cumulative incidence of different ancestries or countries where the hazard ratios, confidence intervals and *P*-values were derived from a Cox proportional-hazards model, adjusted for sex. The reference category of the assessed ancestries or countries was set to European ancestry (EUR) and United States of America (USA), respectively. Affected and unaffected LRRK2 p.Gly2019Ser variant carriers were included in the model and the outcome was age at onset or age at examination with right censoring of the affection status. *N*=Number of individuals, AJ=Ashkenazi Jewish ancestry, EUR=General European ancestry, ISR=Israel, USA=United States of America.

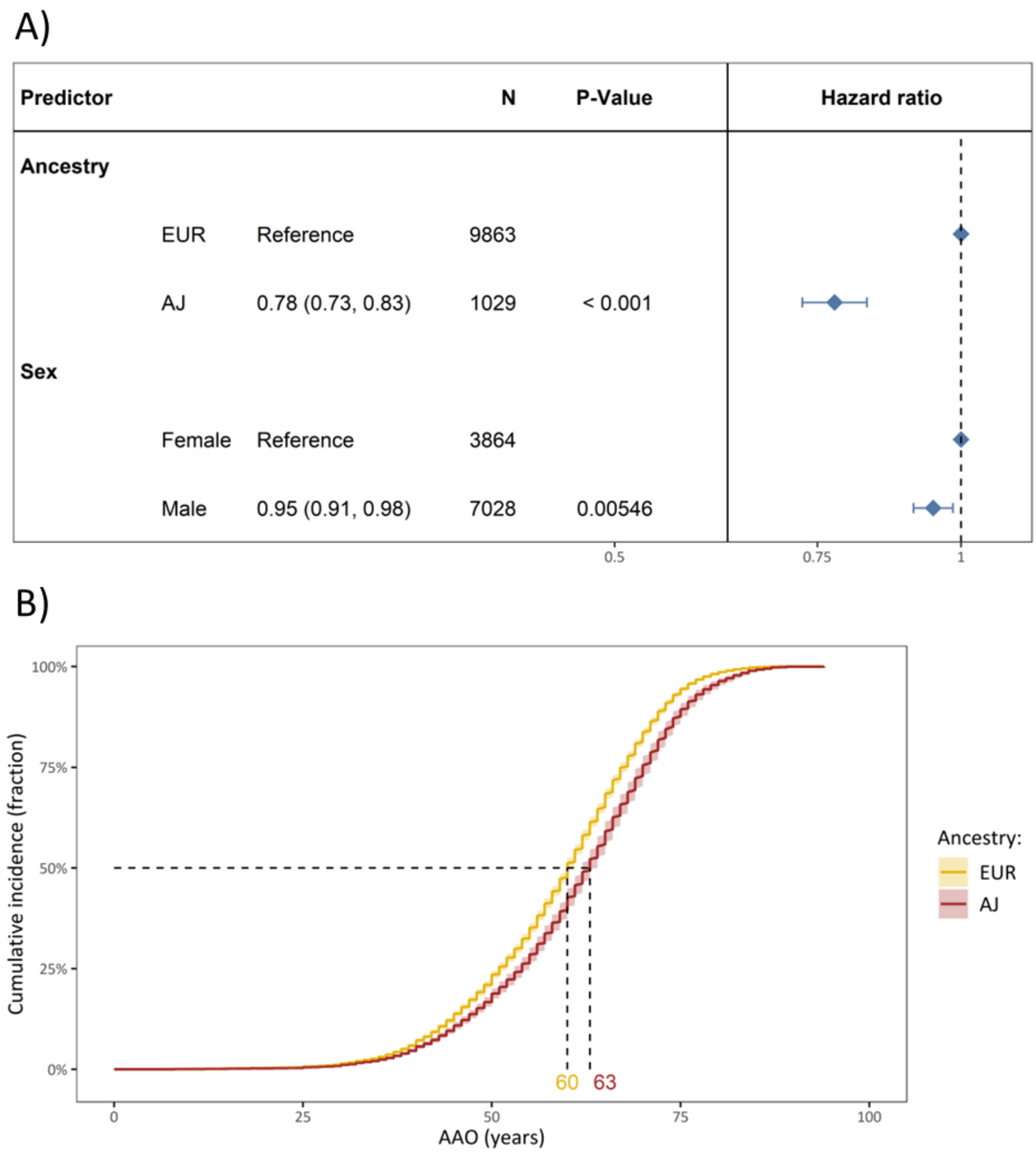


**Supplementary Figure 5. The difference in cumulative incidence of PD patients that do not carry the LRRK2 p.Gly2019Ser from different genetic ancestries. (A)** The forest plot indicates the difference in cumulative incidence of different ancestries where the hazard ratios, confidence intervals and *P*-values were derived from a Cox proportional-hazards model, adjusted for sex. The reference category of the assessed ancestries was set to European ancestry (EUR). (**B**) The Kaplan-Meier plot shows the adjusted curves, visualizing the Cox proportional-hazards model displayed above.

*N*=Number of individuals, AAO=age at onset, PD=Parkinson's disease, AJ=Ashkenazi Jewish ancestry, EUR=General European ancestry.

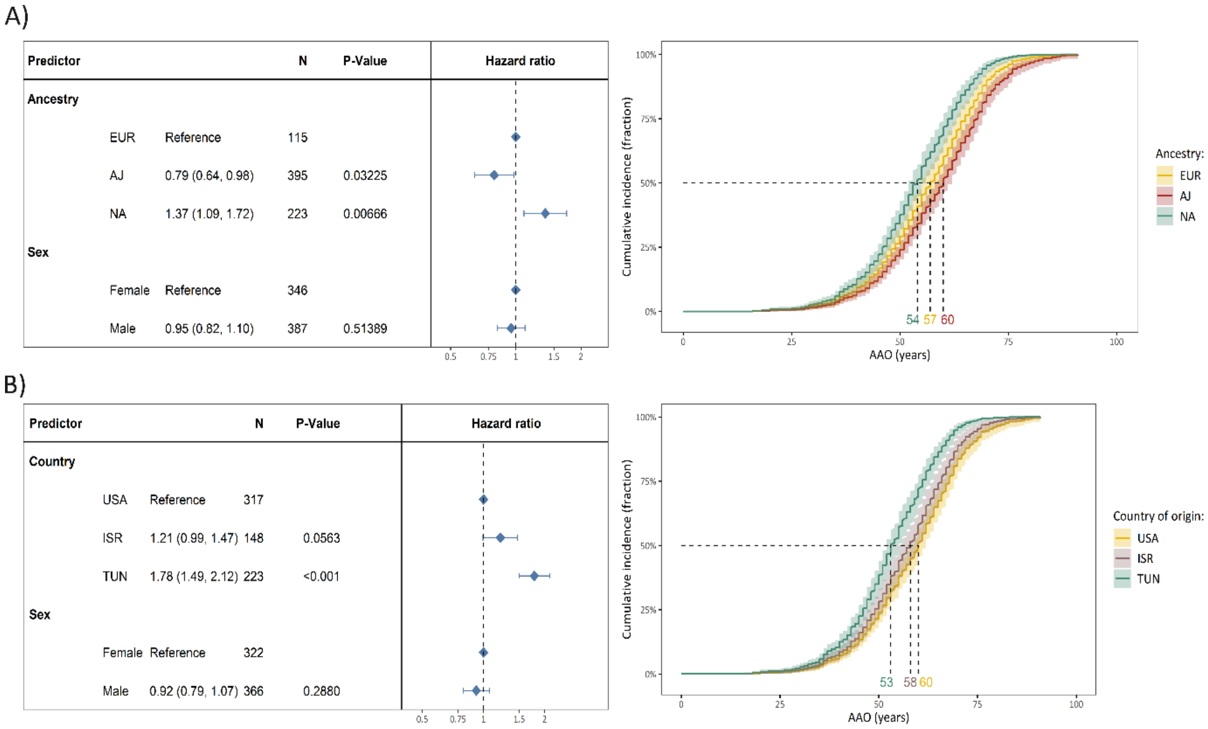


**Supplementary Figure 6. The difference in cumulative incidence of LRRK2 p.Gly2019Ser variant carriers from different genetic ancestries and countries. (A)** The forest plot indicates the difference in cumulative incidence of different ancestries where the hazard ratios, confidence intervals and *P*-values were derived from a Cox proportional-hazards model, adjusted for sex. The reference category of the assessed ancestries was set to European ancestry (EUR). Only affected LRRK2 p.Gly2019Ser variant carriers were included in the model, and the outcome was age at (AAO). The Kaplan-Meier plot shows the adjusted curves, visualizing the Cox proportional-hazards model displayed above. **(B)** The forest plot indicates the difference in cumulative incidence of different countries where the hazard ratios, confidence intervals and P-values were derived from a Cox proportional-hazards model, adjusted for sex. The reference category of the assessed countries was set to the United States of America (USA). Only affected LRRK2 p.Gly2019Ser variant carriers were included in the model, and the outcome was age at (AAO). The Kaplan-Meier plot shows the adjusted curves, visualizing the Cox proportional-hazards model displayed above.

*N*=Number of individuals, AJ=Ashkenazi Jewish ancestry, EUR=General European ancestry, ISR=Israel, USA=United States of America, NA=North African ancestry, TUN=Tunisia.
